# A multivariate spatiotemporal spread model of COVID-19 using ensemble of ConvLSTM networks

**DOI:** 10.1101/2020.04.17.20069898

**Authors:** Swarna kamal Paul, Saikat Jana, Parama Bhaumik

## Abstract

The high R-naught factor of SARS-CoV-2 has created a race against time for mankind and it necessitates rapid containment actions to control the spread. In such scenario short term accurate spatiotemporal predictions can help understanding the dynamics of the spread in a geographic region and identify hotspots. However due to the novelty of the disease there is very little disease specific data generated yet. This poses a difficult problem for machine learning methods to learn a model of the epidemic spread from data. A proposed ensemble of convolutional LSTM based spatiotemporal model can forecast the spread of the epidemic with high resolution and accuracy in a large geographic region. A data preparation method is proposed to convert available spatial causal features into set of 2D images with or without temporal component. The model has been trained with available data for USA and Italy. It achieved 5.57% and 0.3% mean absolute percent error for total number of predicted infection cases in a 5day prediction period for USA and Italy respectively.

## 1. Introduction

Wuhan city in China initially observed an outbreak of Covid-19 disease caused by SARS-CoV-2. Eventually it became a pandemic and more than 200 countries are fighting hard to contain the infection [1]. This has become a unique challenge for mankind as the competition has turned out to be against time due to exponential rate of infection spread. One of the best ways to contain the infection is rapid identification of positive cases and isolation. However due to limited resources random and widespread testing may not be feasible in populous countries. Forecasting regional spread can help identify future hotspots and distribution of infection which would eventually help to take containment measures.

A spatiotemporal epidemic spread model can accommodate both spatial and temporal correlations in data. However, most of the models either require disease specific domain knowledge [11] or are too spatially coarse [18]. Deep learning models can learn the dynamics of epidemic spread with high spatial resolution and high degree of accuracy with minimal initial bias due to its capability of highly nonlinear representation. Deep neural network based spatiotemporal models [4] have already been applied to predict epidemic spread. However, this model is experimented on a small localized region and influence of external factors are ignored. Deep learning models also tend to overfit due to its high representational capability. Due to availability of very limited dataset the problem of overfitting looms large in this case. Thus, modelling of Covid-19 spread in a wide region with high spatial and temporal resolution is challenging.

To address the problem of spatiotemporal prediction of Covid-19 spread in a large geographical region with high resolution, we propose an ensemble of Convolutional LSTM [5] based model to be trained with multilayer temporal geospatial data, transformed as sequence of images. Each layer of the geospatial data corresponds to a causal factor that might influence the spread of the epidemic. An ensemble of models helps reduce variance is errors and overfitting on small training dataset. The data preparation method creates geospatial images of features based on latitudes and longitudes thus avoiding the need of location specific adjacency matrix. We experimented with data of US and Italy and achieved country level mean absolute percent error (MAPE) of 5.57% and 0.3% respectively on forecasting of total infection cases in 5 days period.

The paper is organized as following. In section 2 we conducted a literature review. A brief discussion on modelling the epidemic spread and data preparation method is presented in section 3. In section 4 we explained the ensemble of Convolutional LSTM model and performance measurement metrics. Section 5 is about experimental results. The following section concludes the paper.

## 2. Related Work

Hu et. al. developed a modified stacked autoencoder model of the epidemic spread in China and they claimed to achieve high level of forecasting accuracy [2]. On observing a universality in the epidemic spread in each country, Fanelli and Piazza [3] applied mean-field kinetics of Susceptible-Infected-Recovered/Dead epidemic model to forecast the spread and provided an estimation of peak infections in Italy. Zhan et. al. [8] integrated the intercity migration data in China with Susceptible-Exposed-Infected-Removed model to forecast an estimation of epidemic spread in China. Xi et. al. [4] used deep residual networks to model spatiotemporal characteristics of the spread of influenza and experimented with real dataset of Shenzen city in China. Shi et. al. [5] proposed convolutional LSTM network for spatiotemporal modelling of localized rainfall over a short period of time and used it for rainfall nowcasting. Yuan et. al. [6] used an ensemble of ConvLSTM models to predict road accidents by using heterogeneous multi-layered spatiotemporal data.

## 3. Modelling Covid-19 spread

Disease spread is a complex dynamical system and numerous factors contribute to the dynamics of spread making it non-stationary. Covid19 is no different. Geographical location, weather conditions [7], human mobility [8], population statistics might be some of the impacting factors changing the dynamics of the spread. Epidemic spread is correlated in time as well as spatial dimensions. There is high chance of spreading the infection in spatially co-located geographical regions specially during community spread. Thus, a spatiotemporal model can more accurately capture the dynamics of the spread. However, it may be spatially autocorrelated in a small localized region but not across wide regions. Thus, we divided a large geographic region into relatively smaller grids and trained our model with samples drawn from local distribution of infection cases.

The goal of the model is to forecast new cases of infection on daily basis in different regions across a country which can be added up to calculate total cases of infection. The model can be trained with historical time series data of new cases of infection in different regions along with some external spatial and/or spatiotemporal features. During prediction, the model is fed with temporal sequence of distribution of new infection cases in a region along with other external spatiotemporal features and the model in turn forecasts the distribution of new infection cases in that region.

### 3.1. Data Preparation

All the observations in the dataset is mapped to a spatial region bounded by predefined latitude and longitude. The spatial region may represent a section of a single country or multiple countries. The region is geospatially divided in M x N grids of equal sizes bounded by calculated latitudes and longitudes. It is assumed that disease spread in locations within a grid is spatially autocorrelated due to its smaller size compared to the whole region. Fig. 1a illustrates a grid bounded by latitudes and longitudes. The box represented by the dotted line is called as frame. The frames have overlapping areas in all 4 direction. The overlap allows flow of spatial influence from neighbouring grids. Each frame is in turn divided into L x L pixels which includes the overlapping area. Each pixel represents a bounded area in geospatial region. The values in each pixel is mapped to certain feature in the bounded geospatial region. Separate frame matrices are constructed for each feature and concatenated through channels. For example, new infection count and population are two features and they represent two separate L x L matrices in a frame concatenated across a third axis. Each pixel in the infection count matrix contains the count of new infections (ΔI) in the pixel area in a day. Infection count is distributed both in spatial and temporal dimensions. To reduce the variance, the infection count in a pixel is log transformed and normalized in 0-1 scale. Considering R0 factor of Covid-19 between 2 and 3 it is calculated that 60% of the population (P) in an area needs to get infected to attain herd immunity and reduce further spreading [20]. Similar to the SIR model [14], the total population P is compartmentalized into susceptible (S) and infected/recovered/deceased (I) group. Susceptible population at any day is calculated as 0.6P−I.In a single day new infection count cannot exceed number of susceptible populations. Thus, a pixel value is calculated as In(ΔI +1)/In (S+2). Total population is distributed spatially in similar fashion and it is assumed time invariant within a short interval. Pixel value of population matrix is calculated as In(0.6P+1)/In (max(0.6P)). Each frame is represented as tensor of dimension T x L x L x C, where T is the total time span and C is number of channels or features. As shown in Fig. 1b each training sample in a frame is generated by sliding a time window size of W+1 by 1, leaving behind a test case sample of time window size of W′ in the most recent period. Number of training samples in a frame can be calculated as T−W′−W −1. Thus, total number of training samples *S*_*train*_ for all frames can be calculated as S_train_=(T−W′−W −1) *M*N.

**Fig. 1.**
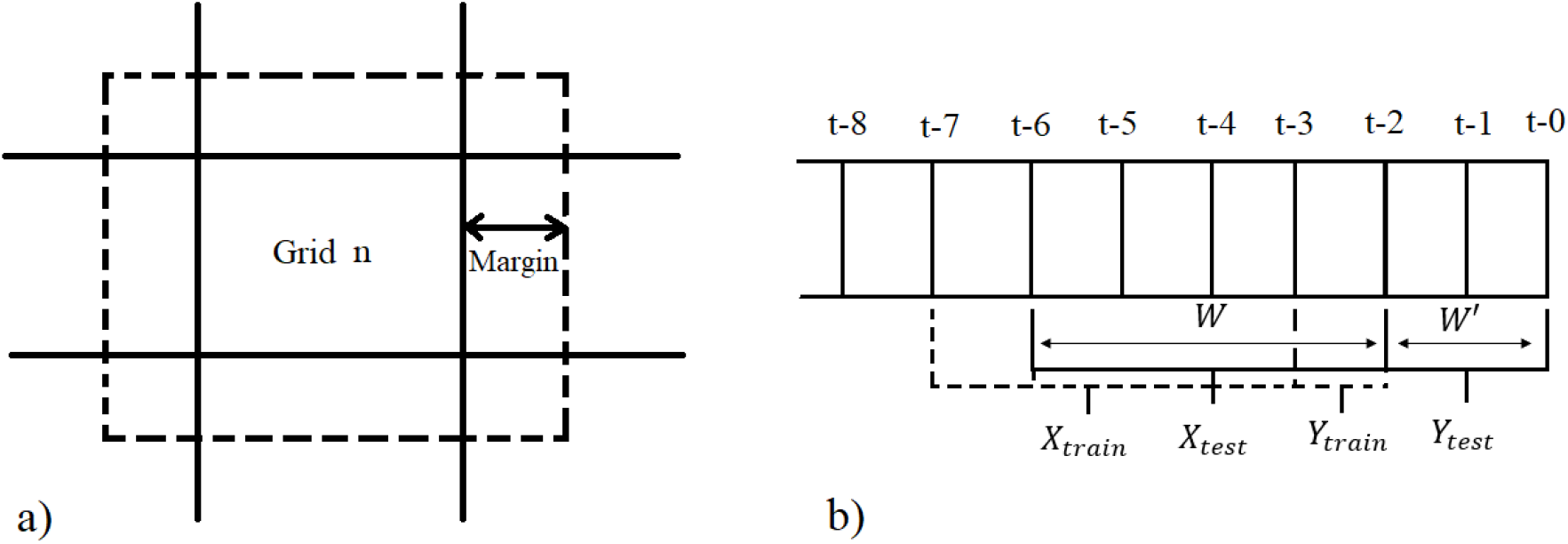
a) Illustration of overlapping frames obtained by spatially dividing a geographical region. The bold lines represent latitudes and longitudes which separates the grids. The box with dotted line represents the overlapping frame which represented as an image for training the model. Each grid is divided in certain number of pixels. The margin refers to number of pixels of overlapping region. b) Illustration of sequence of images in a frame. t-0 is the most recent image. X_train_, Y_train_ are the training samples and X_test_, Y_test_ are testing samples.

The forecasting problem is framed as supervised learning problem. Given a sequence of observed matrices of spatial data as images *X*_1_, *X*_2_…*X*_*t*_ the final objective of the model is to predict the next image *X*_*t*+1_. The training samples are divided into input sequences and output sequences each of length W. The input sequence is time lagged images of output sequence. The model forecasts the normalized log transformed new infections count in each pixel in an image for each timestep. Thus, the output image consists of only 1 channel. The input training dataset (X_train_) can be represented as a tensor of size S_train_⨯W⨯L⨯L⨯Cand the output dataset (Y_train_) as S_train_⨯W⨯L⨯L⨯1. For training, the input sequences are selected from all frames having non-zero total infection count. Fig. 1b illustrates the sequence of images in a frame. The image t-7 to t-3 represents an input training sequence (X_train_) of length W. The output image sequence (Y_train_) for this training sample is t-6 to t-2. Other training samples are generated by sliding the window W backwards in time by 1. The most recent images t-0 and t-1 represents the test output images (Y_test_) and immediate sequence of images t-6 to t-2 is the test input sample (X_test_). The test set X_test_ is represented by a tensor of size (M*N)⨯W⨯L⨯L⨯Cand Y_test_ by (M*N)⨯W′⨯L⨯L⨯1.

## 4. Ensemble of Convolutional LSTM models

Recurrent neural networks (RNN) are a class of artificial neural networks with nodes having feedback connections thereby allowing it to learn patterns in variable length temporal sequences. However, it becomes difficult to learn long term dependencies for traditional RNN due to vanishing gradient problem [9]. LSTMs [10] solve the problem of learning long term dependencies by introducing a specialized memory cells as recurrent unit. The cells can selectively remember and forget long term information in its cell state through some control gates. In convolutional LSTM [5] a convolution operator is added in state to state and input to state transition. All inputs, outputs and hidden states are represented by 3D tensors having 2 spatial dimensions and 1 temporal dimension. This allows the model to capture spatial correlation along with the temporal one. In our model we configured multichannel input such that distinct features can be passed through different channels. Multiple convolutional LSTM layers are stacked sequentially to form a network with high representational capability. The network terminates with a 3D convolutional layer having one filter. This layer constructs a single channel output image as the next frame prediction.

A single model may be prone to overfitting on training dataset and loose stability in terms of prediction made. Creating an ensemble of diverse models intended to solve the same task and combining the predictions made by them typically improves test accuracy and stability [12]. We used bagging or bootstrap aggregation [13] to create an ensemble of models. 60% random samples are drawn with replacement from the original training dataset and an ensemble of five models are trained individually. During prediction the output of each of the models are weighted as per following equation.

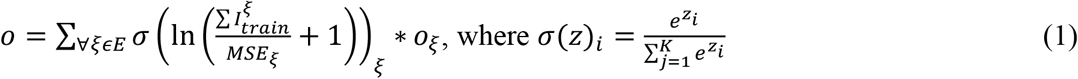

*o* is output of the ensemble, *o*_*ξ*_ is output of the model *ξ, E* is set of all models in the ensemble, 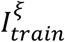 is number of infected patients in the training samples of model *ξ, MSE*_*ξ*_ is mean squared error of model *ξ* on validation dataset and *σ* is softmax function. The weights are proportional to the amount of positive cases used for training the model and inversely proportional to the validation error.

During testing the model is given a sequence of most recent frames as input and the next frame is predicted. The predicted frame is temporally appended to the input sequence of frames and fed to the model again to obtain the next predicted frame. This continues until required number of future frames are predicted. The accuracy of a model is tested with the metric “mean absolute percent error” (MAPE) and Kullback-Liebler (KL) divergence [17]. The pixel values are transformed to ΔI and summed up cumulatively to calculate total infection cases I, up till a specific day. MAPE is calculated at pixel level for total infection cases at the end of prediction period and averaged. The pixels with 0 susceptible population count are ignored while calculating MAPE. Pixel MAPE is calculated as per equation 2, where *G*_*p*_is set of all unique pixels in all grids such that the frame for each corresponding grid have non zero total infection count, *W*′ is prediction time period, *W*^″^=*T* −*W*′ is total time period in training set, *p* is a pixel from a set of unique pixels in the total region having non zero actual susceptible population, *p*_*i*_is predicted pixel value on *i*th day, 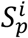 is susceptible population at pixel *p* on *i*th day calculated from *p*_*i*_ and 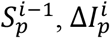 is actual new infection count at pixel *p* on *i*th day and *N*_*p*_is total number of pixels *p* in the region. *Î*_*p*_and *I*_*p*_are total predicted and actual infection cases respectively.

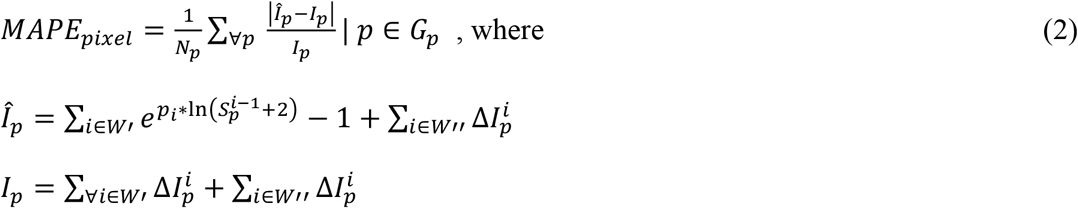

KL divergence at pixel level is calculated for total infection cases at the end of prediction period to measure the dissimilarity of distribution of predicted infection cases with respect to actual. *σ* is softmax function applied after scaling a series in 0 to 1 scale and *P*(*X*) is probability distribution of *X*. Softmax is applied to convert total infection cases as probability distribution across pixels.

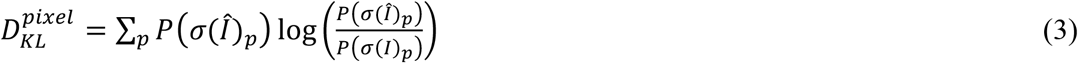

MAPE is also calculated at country level with respect to total predicted infection cases across the region during the prediction period.

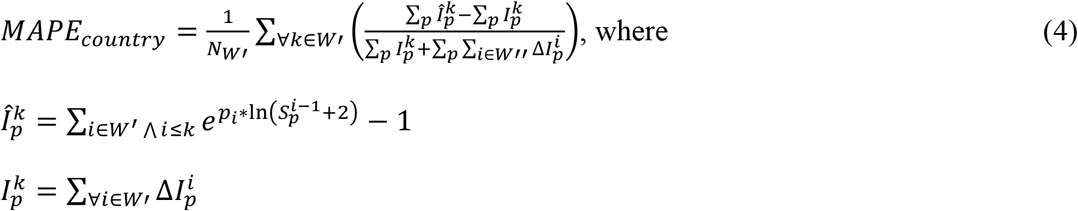

## 5. Experimental Results

Experiments have been carried out to predict the future new infection cases in Italy for a period of 5 days and 10 days and in USA for a period of 5 days and 8 days. Data has been collected from Harvard dataverse [15,16] and [19]. For USA the data collection period is ‘2020-03-09’ to ‘2020-04-08’ and for Italy it is ‘2020-02-05’ to ‘2020-04-10’. Test data period for Italy data is ‘2020-04-01’ to ‘2020-04-10’ and for USA it is ‘2020-04-01’ to ‘2020-04-08’. Fig. 2a shows the region of USA which has been divided into 18×30 grids. The length W of each training input sequence is taken as 10 days. As shown in Fig. 2b, the region of Italy is divided in 7×6 grids. For both the countries the frames containing at least a single Covid-19 infection case will be considered for training and testing the model. The Covid-19 cases are marked in red bubbles in the map. Each frame in turn is divided in 16×16 pixels with an overlap *Margin* of 4 pixel. Training sequences containing at least one positive case are only selected.

**Fig. 2a.**
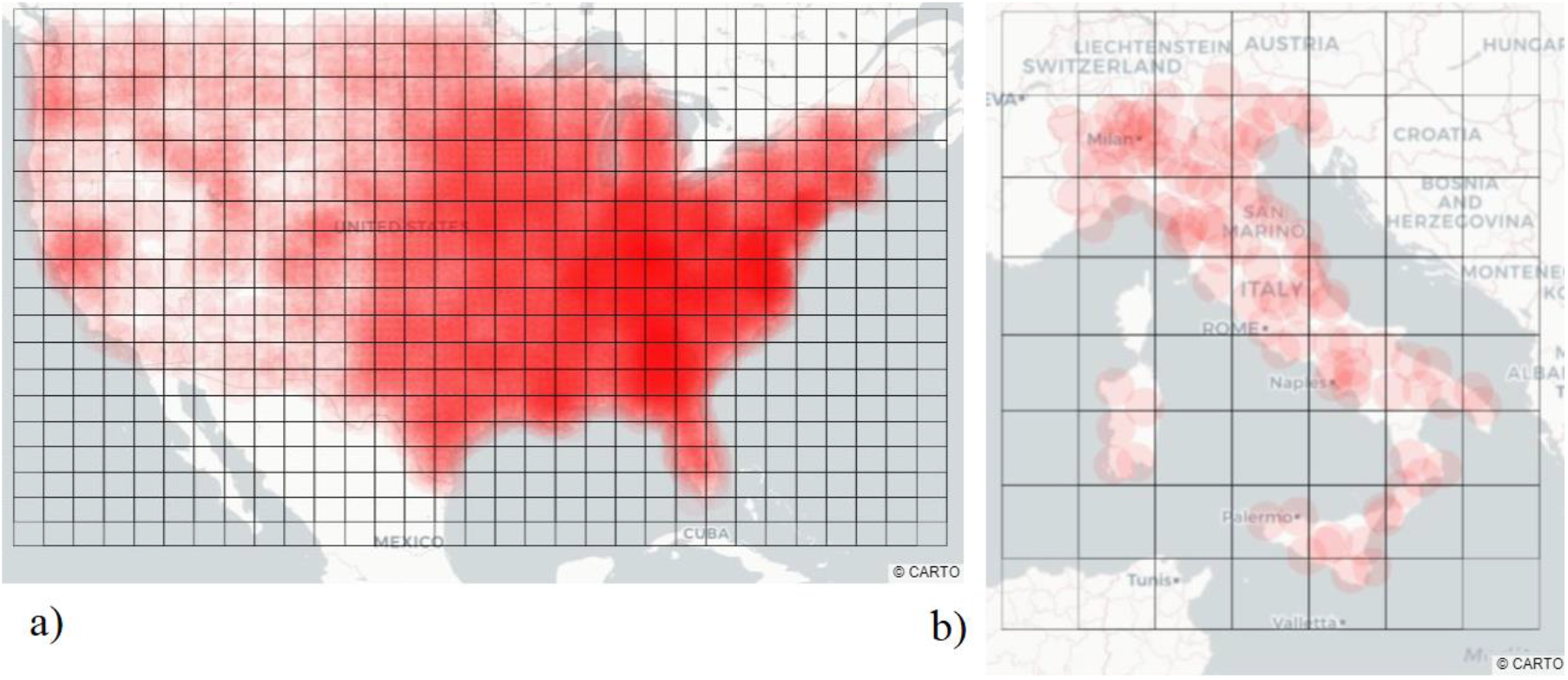
A region of USA divided in 18×30 grids. The red bubbles denote cumulative number of Covid-19 cases. b) A region of Italy divided in 8×7 grids with cumulative Covid-19 cases denoted by red bubbles.

The model consists of ensemble of 5 Convolutional LSTM networks. For Italy each network contains 4 hidden layers with sigmoid activation. The output layer is a Convolutional 3D layer with exponential linear unit as activation. The models are trained for 30 epochs with mean squared error as loss function. Each Conv2D layer has kernel of size 3×3. The Conv3D layer has kernel size 3×3×3. The input and hidden layers have 32 filters. The input layer is configured to take images of size 16×16×2. Each frame in the input sample have 2 channels for 2 features. The first channel contains the normalized log transformed count of new cases per day. The second channel contains the normalized log transformed population in each pixel with no temporal variation. The region in USA is approximately 13 times than Italy and the distribution of Covid-19 cases in USA is geospatially highly skewed. Thus, grids are divided in four equal sections by a latitude and longitude with each section containing 9×15 grids. A set of 4 heterogenous ensembles are trained for each of the 4 sections. The configuration of networks is same as that of Italy except they contain 2 hidden layers and each network is trained for 20 epochs. The implementation has been done on Python and code is available at https://github.com/swarna-kpaul/covid19spatiotemporal.

Table I shows the performance of the models in terms of KL divergence and MAPE. For both USA and Italy, low KL divergences states that the predicted geospatial probability distribution of total infection cases nearly matches with the actual probability distribution. The pixel level MAPE for Italy stays below 30%. For USA in 8-day forecasting period MAPE is 44% as there are many pixels in USA with low total patient count. A slight deviation in the prediction for these pixels shoots up the MAPE. Country level MAPE is low for both Italy and USA.

**Table 1.** KL divergence at pixel level and mean absolute percent error at pixel and country level for Italy and USA

| Country | Pixel KL divergence | Pixel MAPE Total cases | Country MAPE Total cases | Predicted Total cases | Test span in days |
| --- | --- | --- | --- | --- | --- |
| Italy | $5.72 \times 10^{-5}$ | 11.51% | 0.3% | 129665 | 2020-04-01 – 2020-04-05 |
| Italy | 0.00010 | 28.64% | 1.4% | 151803 | 2020-04-01 – 2020-04-10 |
| USA | $3.06 \times 10^{-6}$ | 30.4% | 5.57% | 292032 | 2020-04-01 – 2020-04-05 |
| USA | 0.0014 | 44% | 9% | 368660 | 2020-04-01 – 2020-04-08 |

Fig. 3a and 4a shows predicted vs actual total Covid-19 cases for a period of 8 and 10 days in USA and Italy respectively. For Italy the prediction follows closely with the actual whereas for USA it is little underestimated. Fig. 3b and 4b shows predicted vs actual daily new Covid-19 cases for a period of 8 and 10 days in USA and Italy respectively. Fig. 5a and 5b shows the distribution of total predicted vs actual infection cases in each pixel after 10 day and 8 day in Italy and USA respectively. The predicted distribution closely follows with actual with residuals distributed both on negative and positive side.

**Fig. 3.**
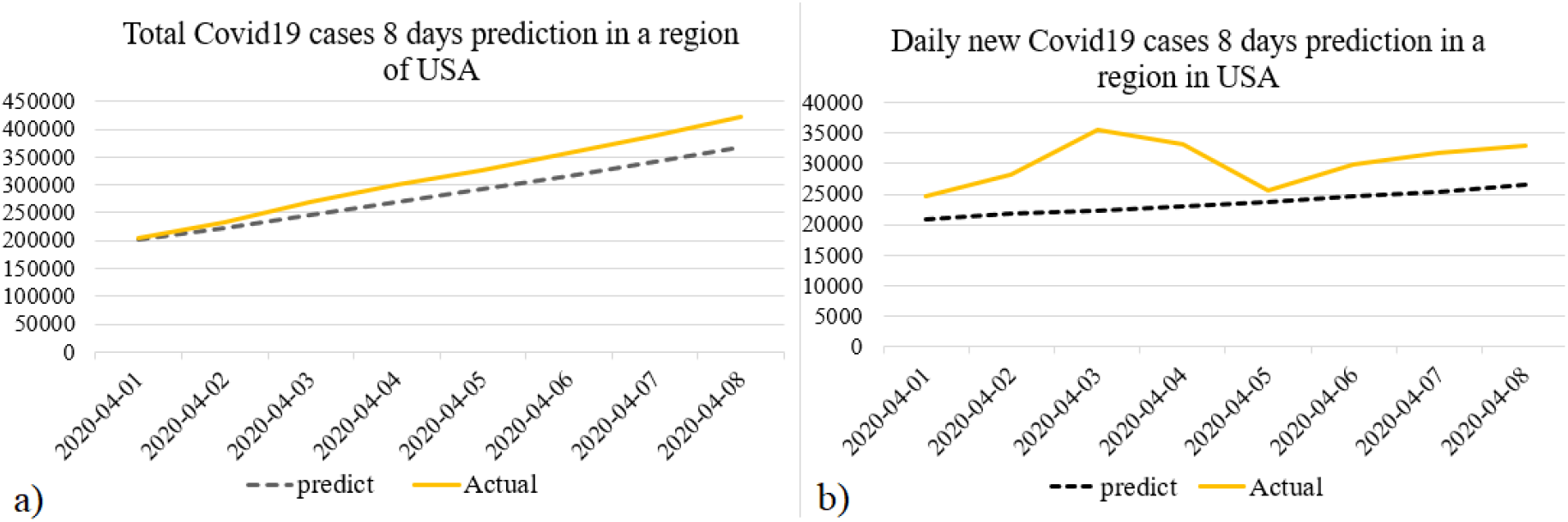
a) Predicted vs Actual total Covid-19 cases for a period of 8 days in a region of USA. b) Predicted vs Actual daily new Covid-19 cases for a period of 8 days in a region of USA

**Fig. 4.**
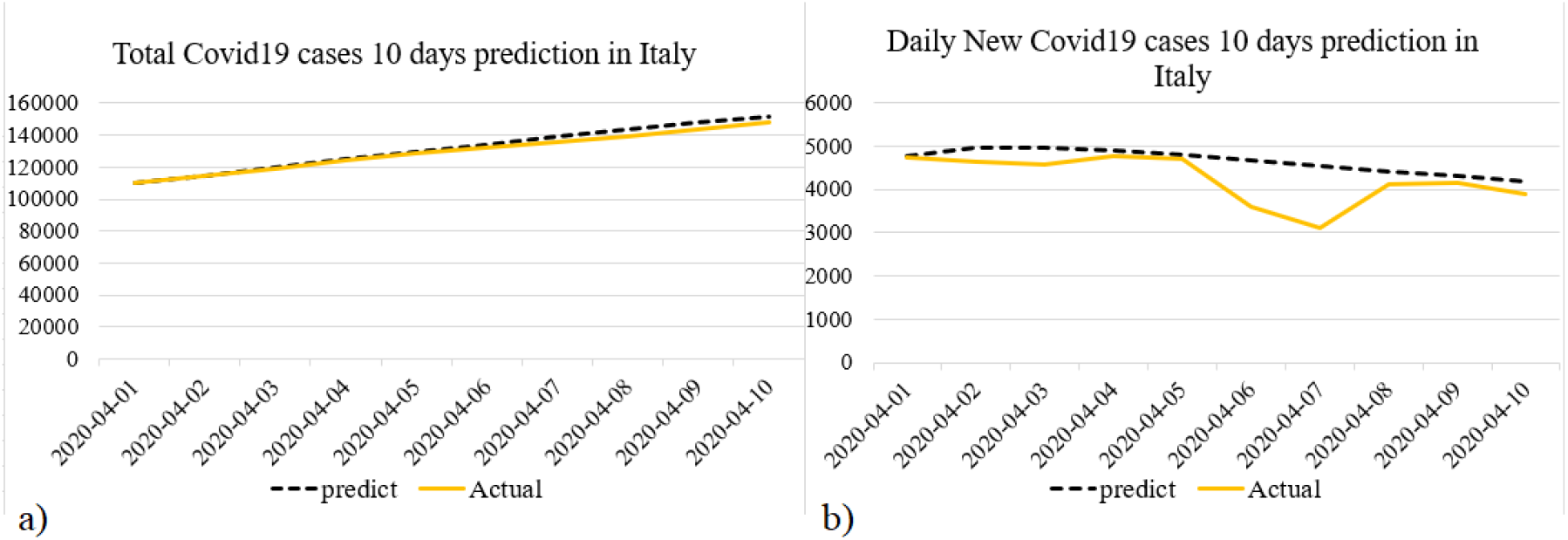
a) Predicted vs Actual total Covid-19 cases for a period of 10 days in a region of Italy. b) Predicted vs Actual daily new Covid-19 cases for a period of 10 days in a region of Italy

**Fig. 5.**
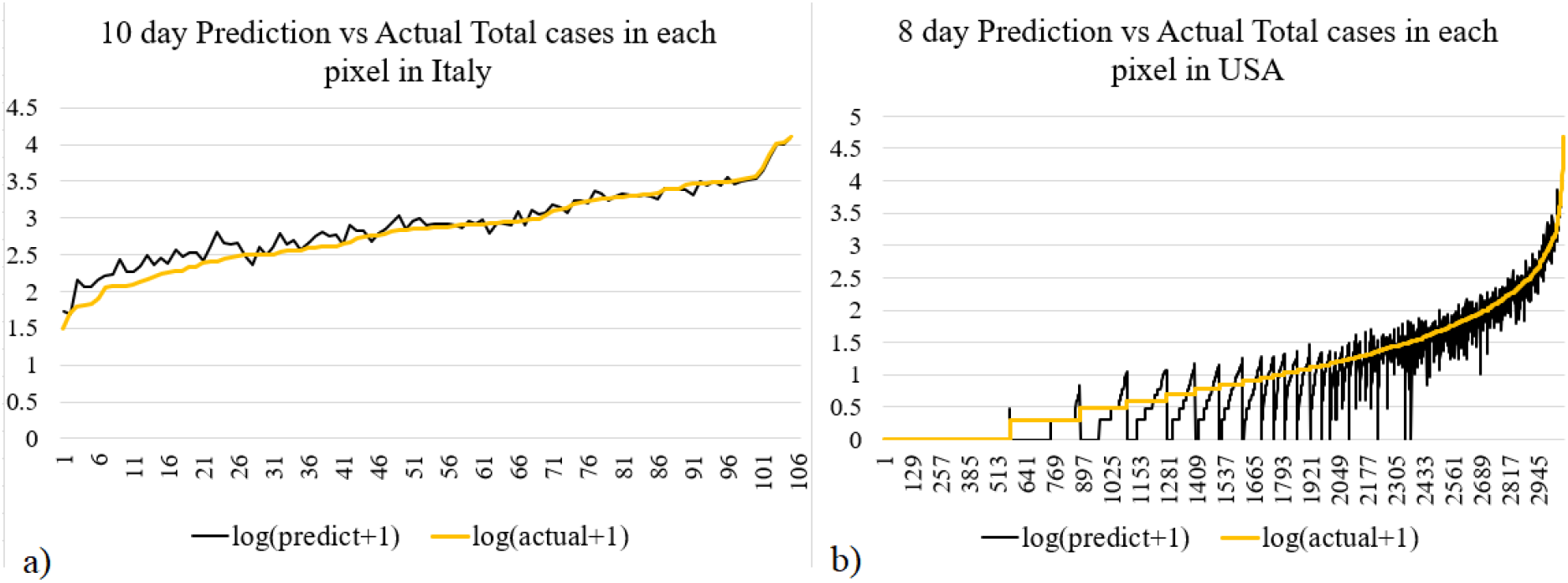
a) Log transformed Predicted vs Actual total Covid-19 cases after a period of 10 days in each pixel in Italy. b) Log transformed Predicted vs Actual total Covid-19 cases after a period of 8 days in each pixel in USA

## 6. Conclusion

An ensemble of Convolutional LSTM based spatiotemporal epidemic spread model has been proposed for short term forecasting of Covid-19 spread. Experiments done on data obtained for USA and Italy reveals high prediction accuracy with high resolution. Since the model has option to fed in any number of external features so we are experimenting with multiple external features that might influence the spread. This might help to find important causal features that are impacting the spread across multiple locations. There are several protected datasets which are not readily available in public domain, like human mobility. We firmly believe such datasets are important causal factors and can improve the spread model if incorporated as an input feature. We are also trying to combine models of multiple countries so that a single ensemble of model can be trained through transfer learning and will eventually help predicting cases across the globe.

## Data Availability

Code is available at github. External data has been taken from dataverse.harvard.edu

## Notes

### Competing Interest Statement

The authors have declared no competing interest.

### Funding Statement

Not applicable.

## References

[1] https://www.who.int/docs/default-source/coronaviruse/situation-reports/20200401-sitrep-72-covid-19.pdf?sfvrsn=3dd8971b_2).

[2] Hu, Z., Ge, Q., Jin, L., & Xiong, M. (2020). “Artificial intelligence forecasting of covid-19 in china”. arXiv preprint 2002.07112.

[3] Fanelli, D., & Piazza, F. (2020). “Analysis and forecast of COVID-19 spreading in China, Italy and France”. Chaos, Solitons & Fractals, 134, 109761.

[4] Xi, G., Yin, L., Li, Y., & Mei, S. (2018, November). “A deep residual network integrating spatial-temporal properties to predict influenza trends at an intra-urban scale”. In Proceedings of the 2nd ACM SIGSPATIAL International Workshop on AI for Geographic Knowledge Discovery (pp. 19–28).

[5] Xingjian, S. H. I., Chen, Z., Wang, H., Yeung, D. Y., Wong, W. K., & Woo, W. C. (2015). “Convolutional LSTM network: A machine learning approach for precipitation nowcasting”. In Advances in neural information processing systems (pp. 802–810).

[6] Yuan, Z., Zhou, X., & Yang, T. (2018, July). “Hetero-convlstm: A deep learning approach to traffic accident prediction on heterogeneous spatio-temporal data”. In Proceedings of the 24th ACM SIGKDD International Conference on Knowledge Discovery & Data Mining (pp. 984–992).

[7] Sajadi, M. M., Habibzadeh, P., Vintzileos, A., Shokouhi, S., Miralles-Wilhelm, F., & Amoroso, A. (2020). “Temperature and latitude analysis to predict potential spread and seasonality for covid-19”. Available at SSRN 3550308.

[8] Zhan, C., Tse, C., Fu, Y., Lai, Z., & Zhang, H. (2020). “Modelling and Prediction of the 2019 Coronavirus Disease Spreading in China Incorporating Human Migration Data”. Available at SSRN 3546051.

[9] Hochreiter, S. (1998). “The vanishing gradient problem during learning recurrent neural nets and problem solutions”. International Journal of Uncertainty, Fuzziness and Knowledge-Based Systems, 6(02), 107–116.

[10] Hochreiter, S., & Schmidhuber, J. (1997). “LSTM can solve hard long time lag problems”. In Advances in neural information processing systems (pp. 473–479).

[11] Neilan, R. M., & Lenhart, S. (2011). “Optimal vaccine distribution in a spatiotemporal epidemic model with an application to rabies and raccoons”. Journal of mathematical analysis and applications, 378(2), 603–619.

[12] Lee, S., Purushwalkam, S., Cogswell, M., Crandall, D., & Batra, D. (2015). “Why M heads are better than one: Training a diverse ensemble of deep networks”. arXiv preprint 1511.06314.

[13] Breiman, L. (1996). “Bagging predictors”. Machine learning, 24(2), 123–140.

[14] Hethcote, H. W. (1976). “Qualitative analyses of communicable disease models”. Mathematical Biosciences, 28(3-4), 335–356.

[15] China Data Lab, 2020, “US COVID-19 Daily Cases with Basemap”, https://doi.org/10.7910/DVN/HIDLTK, Harvard Dataverse, V18, UNF:6:s0u1J15PWisF3mouUiT6Kw== [fileUNF]

[16] Dipartimento della Protezione Civile, 2020, “Coronavirus Disease 2019 (COVID-19) in Italy”, https://doi.org/10.7910/DVN/KDFYZW, Harvard Dataverse, V24, UNF:6:MkKKPk4k39f2yOvf2A///A== [fileUNF]

[17] Kullback, S., & Leibler, R. A. (1951). “On information and sufficiency”. The annals of mathematical statistics, 22(1), 79–86.

[18] Pitzer, V. E., Viboud, C., Simonsen, L., Steiner, C., Panozzo, C. A., Alonso, W. J., … & Grenfell, B. T. (2009). “Demographic variability, vaccination, and the spatiotemporal dynamics of rotavirus epidemics”. Science, 325(5938), 290–294.

[19] https://www.citypopulation.de/en/italy/admin/

[20] Fine, P., Eames, K., & Heymann, D. L. (2011). “Herd immunity: a rough guide”. Clinical infectious diseases, 52(7), 911-916.

